# Serological responses to COVID-19 booster vaccine in England

**DOI:** 10.1101/2021.11.22.21266692

**Authors:** Georgina Ireland, Heather Whitaker, Shamez N Ladhani, Frances Baawuah, Vani Subbarao, Ezra Linley, Lenesha Warrener, Michelle O’Brien, Corinne Whillock, Paul Moss, Mary E Ramsay, Gayatri Amirthalingam, Kevin E Brown

**Affiliations:** Immunisation and Vaccine Preventable Diseases Division, UK Health Security Agency, London, United Kingdom; Statistics, Modelling and Economics Department, UK Health Security Agency, London, United Kingdom; Brondesbury Medical Centre, Kilburn, London, United Kingdom; Sero-Epidemiology Unit, UK Health Security Agency, Manchester, United Kingdom; Virus Reference Department, UK Health Security Agency, London, United Kingdom; Institute of Immunology and Immunotherapy, University of Birmingham, Edgbaston, United Kingdom; Paediatric Infectious Diseases Research Group, St. George’s University of London, London, United Kingdom

**Author notes:** Correspondence: Georgina Ireland. These authors contributed equally (Georgina Ireland, Heather Whitaker). These authors jointly supervised this work (Gayatri Amirthalingam, Kevin E Brown). From Friday 1st October 2021, the UK Health Security Agency (UKHSA) became fully operational. UKHSA takes on the health protection responsibilities of Public Health England (PHE) and incorporates NHS Test & Trace and the Joint Biosecurity Centre (JBC). UKHSA is an executive agency of the Department of Health and Social Care. It is responsible for planning, preventing and responding to public health threats, and providing intellectual, scientific and operational leadership at national and local level, as well as on the global stage.

**Keywords:** COVID-19, COVID-Vaccine, Antibody, Spike Protein, Immunity, Pfizer, AstraZeneca

## Abstract

**Introduction:** There are limited data on immune responses after COVID-19 vaccine boosters in individuals receiving primary immunisation with BNT162b2 (Pfizer-BioNTech) or AZD1222 (AstraZeneca).

**Methods:** A prospective, cohort study to assess SARS-CoV-2 antibody responses before and after booster vaccination with BNT162b2 in adults receiving either (i) two BNT162b2 doses <30 days apart (BNT162b2-control), (ii) two BNT162b2 doses ≥30 days apart (BNT162b2-extended) or (iii) two AZD1222 doses ≥30 days apart (AZD1222-extended) in London, England. SARS-CoV-2 spike protein antibody geometric mean titres (GMTs) before and 2-4 weeks after booster were compared.

**Results:** Of 750 participants, 626 provided serum samples for up to 38 weeks after their second vaccine dose. Antibody GMTs peaked at 2-4 weeks after the second dose, before declining by 68% at 36-38 weeks after dose 2 for BNT162b2-control participants, 85% at 24-29 weeks for BNT162b2-extended participants and 78% at 24-29 weeks for AZD1222-extended participants. Antibody GMTs was highest in BNT162b2-extended participants (942 [95%CI, 797-1113]) than AZD1222-extended (183 [124-268]) participants at 24-29 weeks or BNT162b2-control participants at 36-38 weeks (208; 95%CI, 150-289). At 2-4 weeks after booster, GMTs were significantly higher than after primary vaccination in all three groups: 18,104 (95%CI, 13,911-23,560; n=47) in BNT162b2-control (76.3-fold), 13,980 (11,902-16,421; n=118) in BNT162b2-extended (15.9-fold) and 10,799 (8,510-13,704; n=43) in AZD1222-extended (57.2-fold) participants. BNT162b2-control participants (median:262 days) had a longer interval between primary and booster doses than BNT162b2-extended or AZD1222-extended (both median:186 days) participants.

**Conclusions:** We observed rapid serological responses to boosting with BNT162b2, irrespective of vaccine type or schedule used for primary immunisation, with higher post-booster responses with longer interval between primary immunisation and boosting. Boosters will not only provide additional protection for those at highest risk of severe COVID-19 but also prevent infection and, therefore, interrupt transmission, thereby reducing infections rates in the population. Ongoing surveillance will be important for monitoring the duration of protection after the booster.

## Introduction

Two doses of BNT162b2 (Pfizer-BioNTech/Comirnaty^®^) or AZD1222 (AstraZeneca/Vaxzevria^®^) vaccines provide high levels of protection from COVID-19, hospitalisation and death for at least 4-6 months after vaccination.^1^ Unlike other countries that that offered COVID-19 vaccination according to the authorised 3-4 week interval, the UK recommended an extended 12-week schedule to expedite the rollout of the first dose of vaccine.^1^ Subsequent studies have demonstrated higher antibody levels after the second dose with the extended schedule than the authorised interval, potentially providing better longer-term protection.^1,2^ However, antibody levels and clinical protection wane over time. In England, vaccine effectiveness (VE) against hospitalisation declined to 77.0% and 92.7% beyond 20 weeks post-vaccination and 78.7% and 90.4% against death for AZD1222 and BNT162b2, respectively, with greater waning among older adults and those with underlying comorbidities.^2^ As these groups were the first to be offered COVID-19 vaccination and because of concerns about waning immunity, emergence of the highly-transmissible Delta variant which causes more severe disease and can infect vaccinated individuals,^3,4^ high and sustained community infection rates in the UK, and winter pressures on the national healthcare system, the UK Joint Committee on Vaccination and Immunisation (JCVI) recommended a third dose (booster) with either a single dose of BNT162b2 or a half dose (50µg) of mRNA-1273 (Moderna/Spikevax^®^) for adults aged ≥50 years, individuals aged 16-49 years in clinical risk groups, adult carers and household contacts of immunosuppressed individuals, and frontline health and social care workers, to be offered at least 6 months after their second dose.^5-7^

In England, the UK Health Security Agency (UKHSA) initiated an evaluation of vaccine responses in adults aged ≥50 years who received the BNT162b2 or AZD1222 as part of the national immunisation programme to compare short versus longer interval vaccine schedules and monitor antibody waning over time.^8^ Here, we describe the antibody kinetics after primary immunisation and booster vaccination in adults aged ≥50 years who were vaccinated as of the national COVID-19 immunisation programme.

## Methods

The COVID-19 vacci<u>n</u>e responses after <u>e</u>xtended immunisation sched<u>u</u>les (CONSENUS) cohort has been described previously.^1,8^ CONSENSUS recruited immunocompetent adults aged ≥50 years in January 2021 in London to provide serial blood samples at 0,3,6,9,12,15 and 20 weeks after their first dose of COVID-19 vaccine. As part of the national COVID-19 vaccine roll out, participants received either (i) two BNT162b2 doses <30 days apart (BNT162b2-control), (ii) two BNT162b2 doses ≥30 days apart (BNT162b2-extended) or (iii) two AZD1222 doses ≥30 days apart (AZD1222-extended). Additional blood samples were taken before and after the booster.

Serum samples were tested for nucleoprotein (N) antibodies as a marker of previous SARS-CoV-2 infection (Elecsys Anti-SARS-CoV-2 total antibody assay, Roche Diagnostics, Basel, Switzerland: positive ≥1 COI) and spike (S) protein antibodies, which could be infection-or vaccine-derived (Elecsys Anti-SARS-CoV-2 S total antibody assay, Roche Diagnostics: positive⍰≥⍰0.8 arbitrary units (au)/mL to assess vaccine response). Antibody geometric mean titres (GMTs) were calculated with 95% confidence intervals (CI). Geometric mean ratios (GMR) of responses between timepoints were estimated using a mixed regression model on log responses including a random effect for each participant; separate models were fitted for each vaccine group. The GMR of responses by vaccine type at each post-vaccination timepoint was estimated via regression on log Roche S responses and included age-group and sex. Statistical analyses were performed using STATA v.14.2. Individuals with ≥0.4 (au)/mL on the Roche N assay were considered to have had prior SARS-CoV-2 infection.

## Results

### Antibody kinetics following primary course

Of the 750 recruited participants, 626 provided serum samples for up to 38 weeks after their second dose of vaccine (**Table 1**). Antibody GMTs peaked at 2-4 weeks after the second dose and then declined for all three vaccine schedule groups for all subsequent sampling points (**Table 2**). Antibody GMTs declined by 68% at 36-38 weeks after dose 2 for BNT162b2-control participants, 85% at 24-29 weeks for BNT162b2-extended participants and 78% at 24-29 weeks for AZD1222-extended participants. Antibody GMTs at 24-29 weeks remained higher in the BNT162b2-extended group than in AZD1222-extended participants (942 [95%CI, 797-1113] vs 183 [124-268]; p<0.001), while the latter was similar to BNT162b2-control participants at 36-38 weeks (208 [150-289]).

**Table 1:**
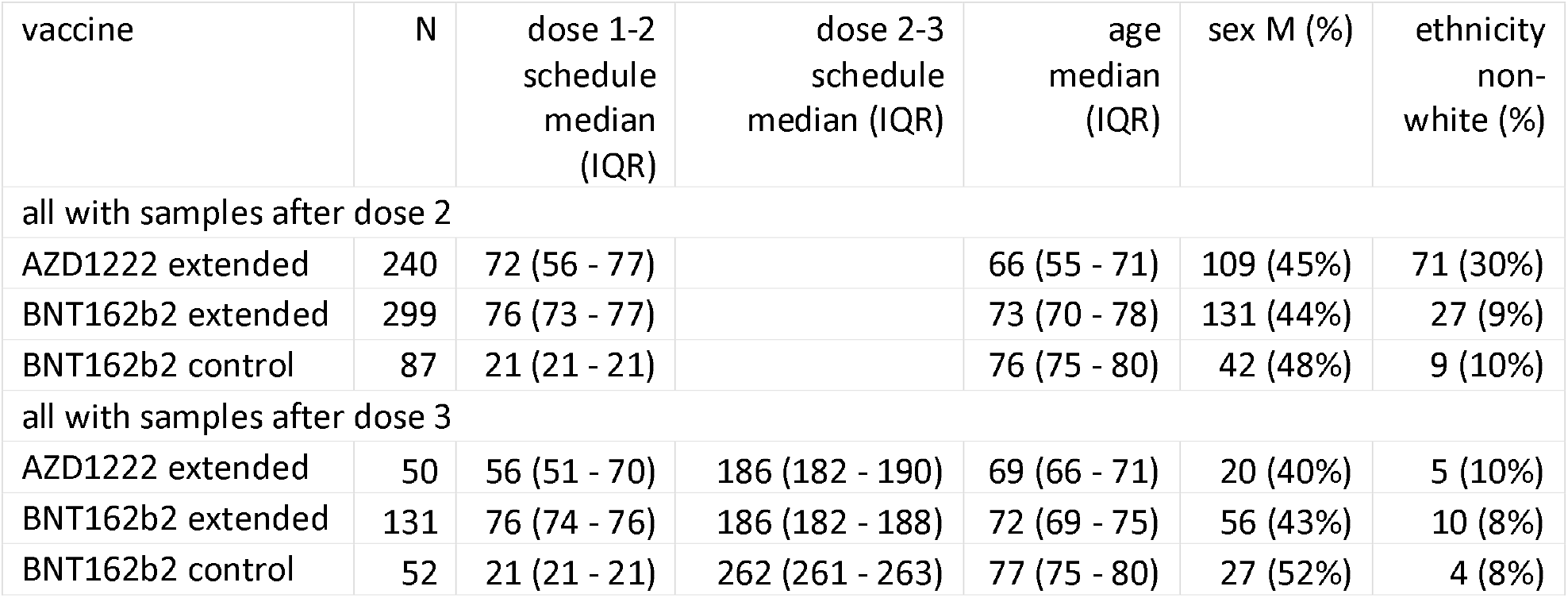
Characteristics of CONSENSUS participants with samples after dose 2 of the primary vaccination and booster vaccine.

**Table 2:**
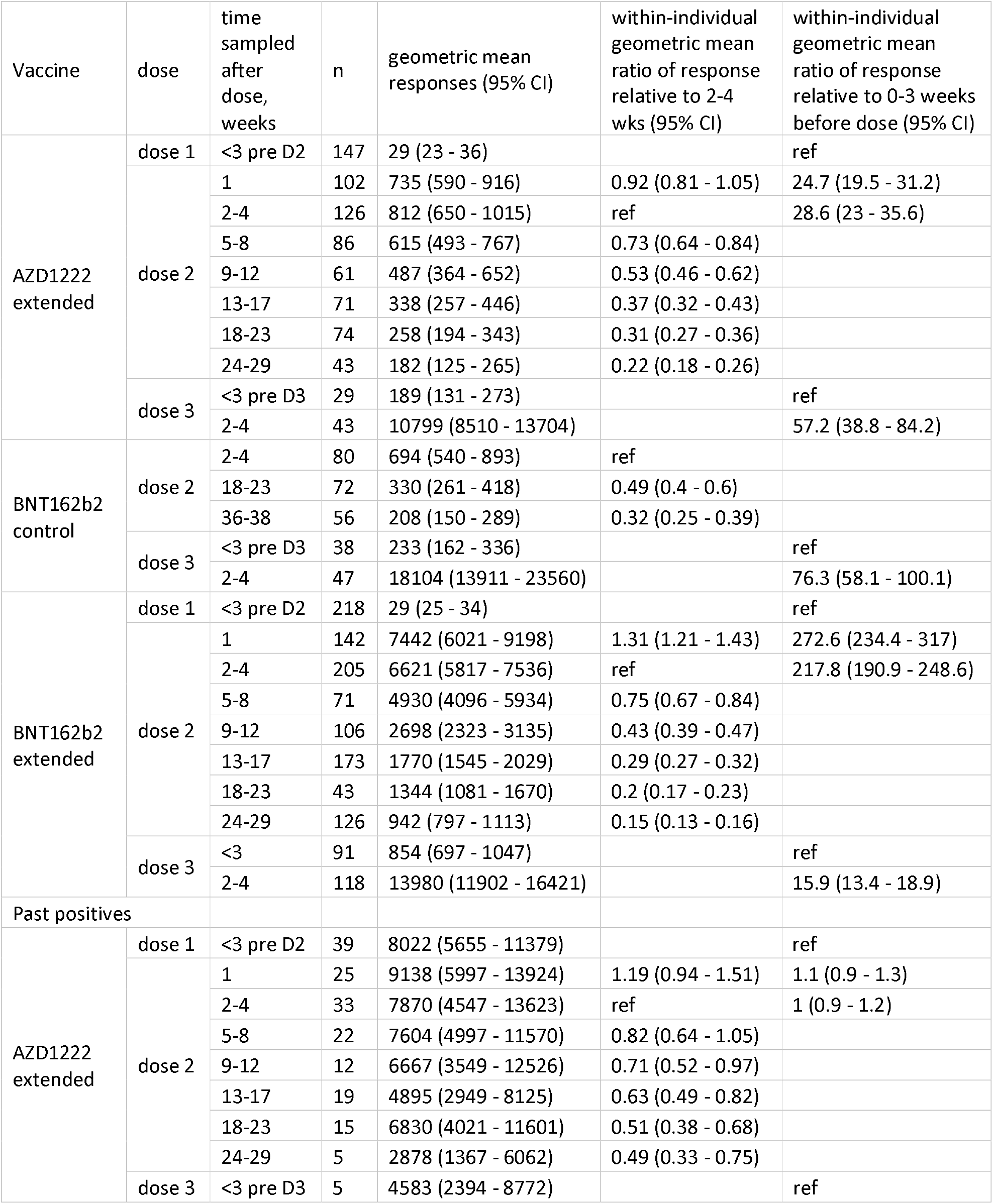
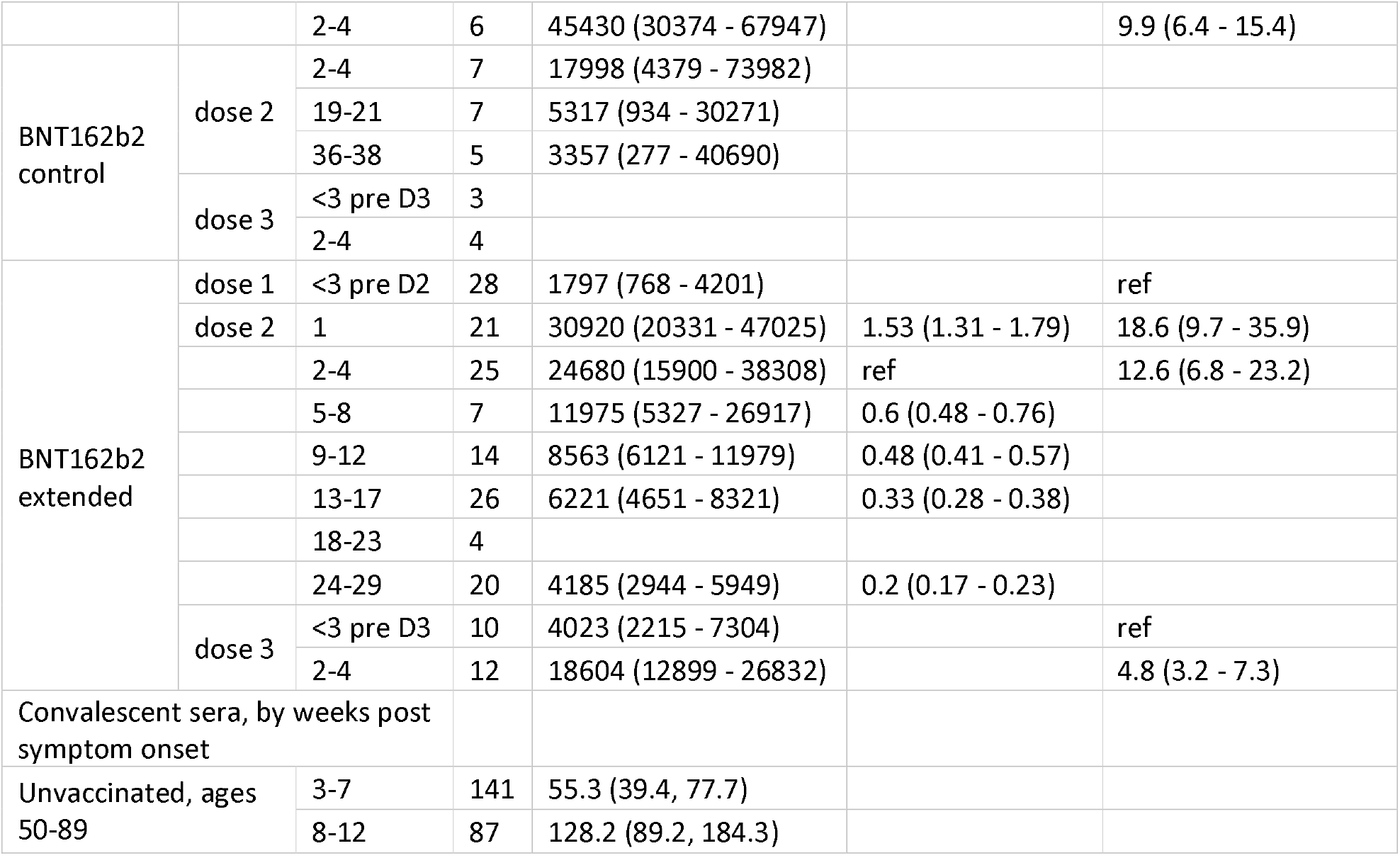
Geometric mean response and geometric mean ratio of response of CONSENSUS participants before and after dose 2 of the primary vaccination and booster vaccine for AZD1222 extended, BNT162b2 control and BNT162b2 extended participants. *Geometric mean response was not calculated for categories with <5 people within them*

Regardless of primary vaccination type or schedule, antibody GMTs at all timepoints after dose 2 were greater in previously-infected participants. In AZD1222-extended participants, however, the decline in GMTs was smaller at 18-23 weeks in previously-infected than in uninfected participants (49% vs 68%). By comparison, a similar decline was observed for previously-infected compared to uninfected BNT162b2-extended participants at 24-29 weeks (80% vs 85%).

### Post-booster response

Serum samples were available for 52 BNT162b2-control, 131 BNT162b2-extended and 50 AZD1222-extended participants 2-4 weeks after the BNT162b2 booster. The boosted AZD1222-extended participants had a shorter interval between primary doses than all AZD1222-extended participants in the evaluation (51 days vs 72 days), while BNT162b2-control participants (median:262 days) had a longer interval between primary and booster doses than BNT162b2-extended or AZD1222-extended participants (both median:186 days).

Antibody GMTs at 2-4 weeks were highest in BNT162b2-control (18,104, 95%CI: 13,9113-23,560), followed by BNT162b2-extended (13,980, 95%CI: 11,902-16,421) and AZD1222-extended (10,799, 95%CI: 8,510-13,704) (**Table 2**). GMTs in BNT162b2-control were greater than for AZD1222-extended participants (p=0.01). The largest post-booster increase in GMTs was in BNT162b2-control participants (76.3-fold), followed by AZD1222-extended (57.2-fold) and BNT162b2-extended (15.9-fold) (Table 2). Booster responses were not affected by age (all p<0.05) but higher in females (p=0.008). Sufficient post-booster information was available for previously-infected AZD1222-extended and BNT162b2-extended participants, where geometric mean responses increased 9.9-fold (to 45,430, 95%CI: 30,374-67,947) and 4.9-fold (to 18,604, 95%CI: 12,899-26,832) respectively.

## Discussion

These early data show a rapid serological response to boosting with BNT162b2, with significantly higher antibody responses than after the second dose. Importantly, our cohort consists primarily of older adults who have a higher risk of severe COVID-19 and are, therefore, most likely to benefit from vaccination. Early data from Israel and England demonstrate significantly better protection against severe COVID-19, hospitalisation and death after booster vaccination.^2,9^ In England, 14 days after boosting with BNT162b2 among ≥50 year-olds, VE was similar in individuals who had received only 2 primary doses of BNT162b2 (87.4, 95%CI 84.9-89.4) and AZD1222 (84.4, 95%CI 82.8-85.8) at least 140 days previously, although the analysis did not separate by schedule.^2^ The higher post-booster GMTs in BNT162b2-control participants, who were the first to be vaccinated, is likely due to the extended interval between primary and booster vaccines, allowing more time for enhancing immune memory and greater waning of antibodies, both of which are likely to enhance post-booster responses.

In conclusion, we observed very high antibody responses following a BNT162b2 booster, irrespective of vaccine or schedule used for primary immunisation. A longer interval between primary immunisation and the booster provides higher post-booster antibody responses and, potentially, longer lasting protection. Decisions on timing of booster doses should take account of the current and predicted epidemiological context to ensure the most vulnerable groups are optimally protected during heightened periods of transmission. Whilst the rates of severe disease, hospitalisations and deaths remain low in individuals receiving primary immunisation only, the booster programme will provide additional protection to those at highest risk of severe COVID-19 and help reduce infection rates across the population.^10^ Ongoing surveillance will be important for monitoring duration of protection offered by booster doses and any need for additional doses in the future.

## Data Availability

Applications for relevant anonymised data should be submitted to the UK Health Security Agency Office for Data Release.

https://www.gov.uk/government/publications/accessing-public-health-england-data/about-the-phe-odr-and-accessing-data

## Acknowledgments

We would like to thank Dorothy Blundell, Dr Caroline Sayer and the team at Haverstock Healthcare GP Federation, and the whole CONSESUS team at PHE including those within the Virus Reference Department at Colindale who assisted with the laboratory testing.

## Author Contributions

SL, FB, PM, MR, KB, GA conceived and designed the study; LW and EL supervised the laboratory testing; SS, CW, MO’B and FB co-ordinated the patient enrolment. HW performed the statistical analysis and GI, GA, SL, and KB wrote the manuscript. All authors read and approved the submission.

---

**Figure 1:**
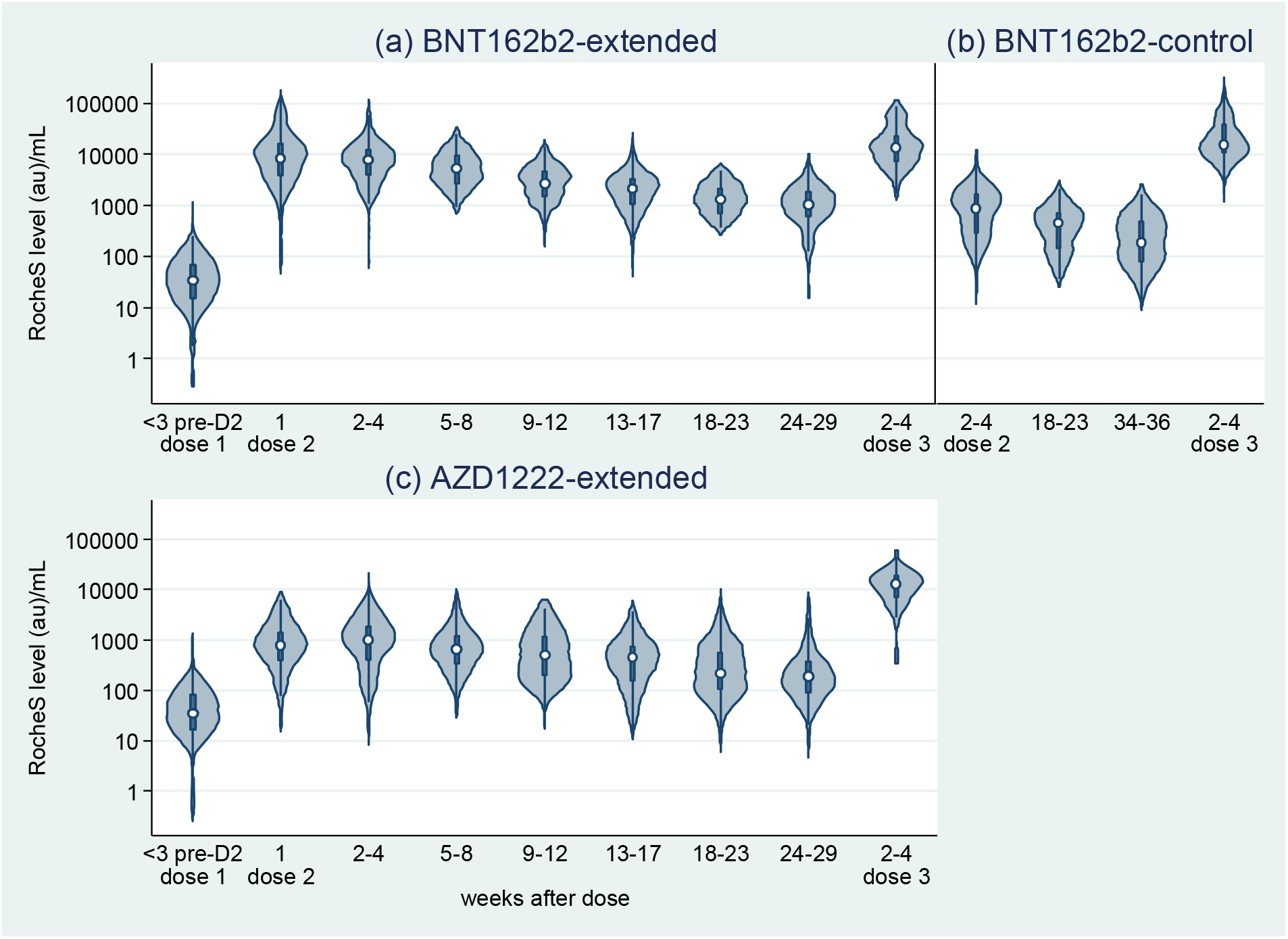
Violin plots, S antibody responses <3 weeks before the 2^nd^ and booster dose of COVID-19 vaccine in previously uninfected individuals whose primary vaccination type was AZD1222-extended schedule, by interval between doses, BNT162b2-extended schedule, by interval between doses and BNT162b2-control schedule.

